# Vascular-Augmented Two-Compartment Fitting Improves Model Performance for Intermittent Myocardial T1 Mapping

**DOI:** 10.64898/2025.12.16.25342336

**Authors:** Yasutoshi Ohta, Tomoro Morikawa, Tatsuya Nishii, Yoshiaki Morita, Tetsuya Fukuda

## Abstract

**Objectives:** Conventional gadolinium-enhanced cardiac magnetic resonance imaging (MRI) typically evaluates myocardial tissues at a single post-contrast time point. In contrast, dynamic T1 mapping enables the estimation of contrast agent concentrations and subsequent pharmacokinetic modeling. This study compared a normal composite two-compartment model incorporating myocardial vascular components with the conventional Brix model.

**Materials and Methods:** This retrospective study included 107 participants who underwent dynamic T1 mapping at 2, 5, 9, and 15 min after contrast administration. Exclusion criteria included contraindications to MR imaging, acute coronary syndrome, pregnancy, an estimated glomerular filtration rate < 30 mL/min/1.73 m^2^, claustrophobia, and known allergy to gadolinium-based contrast medium. Contrast agent concentrations derived from MOLLI-based T1 maps were fitted using the Brix and composite pharmacokinetic models. Model performance was assessed using the residual sum of squares (RSS), Akaike information criterion (AIC), and Bayesian information criterion (BIC). The myocardial blood fraction estimated by the composite model was compared with the extracellular volume (ECV).

**Results:** The composite model exhibited significantly lower RSS, AIC, and BIC values than the Brix model (all p < 0.001). Absolute parameter estimation errors were reduced across all time points. The estimated myocardial blood fraction averaged 35.0% and demonstrated a positive correlation with the ECV (r = 0.61, p < 0.001).

**Conclusions:** In myocardial pharmacokinetic analysis using dynamic T1 maps, the composite model achieved superior fitting performance compared with the Brix model. Explicit incorporation of vascular kinetics improves the longitudinal characterization of contrast behavior and enhances quantitative assessment of myocardial tissue properties.

## Introduction

In myocardial tissue characterization using gadolinium-based contrast agents, assessments are typically performed at a single time point using either late gadolinium enhancement (LGE) imaging or post-contrast T1 mapping to calculate the extracellular volume fraction (ECV). Although these methods provide static evaluations, dynamic (longitudinal) assessments are generally limited to a short temporal window during first-pass perfusion imaging, which is primarily used for ischemia assessment [1].

Dynamic contrast-enhanced (DCE) imaging has been used in oncology to assess tissue perfusion and contrast washout kinetics, enabling the evaluation of tumor subtypes and monitoring of treatment responses [2–4]. Similar temporal imaging approaches have also been applied to the myocardium, with some studies demonstrating correlations between delayed-phase contrast kinetics and myocardial histology [5–8]. However, the clinical implementation of myocardial DCE-magnetic resonance imaging (MRI) remains limited because it requires prolonged continuous image acquisition, making integration into existing cardiac MRI protocols—particularly those already including conventional LGE imaging—challenging without substantially increasing overall scan time.

To address this limitation, a novel approach has been proposed, wherein serial post-contrast T1 maps are acquired intermittently in conjunction with routine LGE sequences. Contrast kinetics can be estimated over time using pharmacokinetic (PK) modeling by converting these T1 values into contrast agent concentrations [9]. This method enables time-resolved myocardial tissue characterization without continuous image acquisition. A commonly employed model in this context is the two-compartment model [10, 11], which offers several advantages, including a limited number of parameters, independence from arterial input function estimation, and compatibility with standard cardiovascular MR protocols.

However, this model may have limitations when applied to organs with substantial vascular beds. Myocardial capillaries are estimated to occupy approximately 20% of myocardial volume during mid-diastole [12], which could compromise the assumptions and accuracy of the conventional two-compartment model. Therefore, we hypothesized that extending the model by incorporating an additional vascular compartment—representing the extracellular vascular space with contrast dynamics similar to those of the blood pool—could improve model fit and enhance precision of the myocardial contrast kinetic analysis.

## Materials and methods

### Participants

This single-center study was a retrospective analysis of data derived from a prospective study (approval no. R20086). Written informed consent was obtained from all participants prior to cardiac MRI. Between December 2022 and December 2023, 107 consecutive patients who underwent dynamic T1 mapping were included in this study. Cardiac MRI was performed to evaluate myocardial properties in patients with suspected cardiomyopathy. Exclusion criteria included contraindications to MR imaging, acute coronary syndrome, pregnancy, an estimated glomerular filtration rate (eGFR) < 30 mL/min/1.73 m^2^, claustrophobia, and known allergy to gadolinium-based contrast medium (CM).

### Image acquisition

Cardiovascular MR examinations were conducted at our institution utilizing a 3T MRI system (MAGNETOM Vida; Siemens Healthcare, Erlangen, Germany) equipped with a 30-channel body coil. MR images were acquired using cine imaging across all cardiac axis views; native T1 mapping; resting perfusion; dynamic T1 mapping at 2, 5, 9, and 15 min following CM administration; and inversion recovery LGE (IR-LGE) true-FISP imaging at 10 min after contrast injection (Figure 2). To ensure compatibility between our imaging and existing perfusion protocols, CM was administered in two split boluses. A dose of 0.05 mmol/kg of CM (Gadobutrol; Gadovist®, Bayer-Yakuhin, Osaka, Japan; injection rate, 5 mL/s; followed by a 10-mL saline flush) was injected for each perfusion imaging session. The interval between the perfusion scans was approximately 100 s. This split-infusion approach minimized the risk of signal saturation in the blood pool during early post-contrast T1 mapping.

Native and post-contrast T1 mapping was performed using the Modified Look-Locker Inversion Recovery (MOLLI) sequence. The following acquisition parameters were applied: repetition time/echo time/flip angle, 3.8 ms/1.2 ms/35°; slice thickness, 5 mm; voxel size, 1.4 × 1.4 × 5.0 mm^3^; number of excitations, 1; field of view, 360 × 306; matrix, 256 × 144; and parallel processing factor, 2 [13]. Three short-axis images (basal, mid-, and apical) were obtained for each acquisition time. T1 maps were generated using inline pixel-wise curve fitting with a three-parameter MOLLI signal model incorporating motion collection and an elastic registration algorithm [14, 15].

### Estimation of tracer concentration

Dynamic post-contrast T1 maps were non-rigidly co-registered to the native T1 map using a commercial software (CVI42, Circle Cardiovascular Imaging). The pixel-wise contrast concentration (c, mM/L) was calculated as follows:

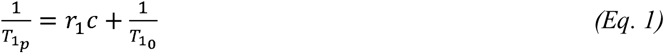

where T_1_ is the post-contrast T1, T_1_ is the native T1, and r_1_ is the relaxivity value of gadobutrol. The r_1_ value (5.0) was used for Gadobutrol at 3.0T [16, 17].

### Pixel-wise PK analysis

#### Brix model

A two-compartment model (modified from Brix and Hoffmann) [10, 11] was used to describe myocardial contrast kinetics. The time after contrast administration was extracted from the Digital Imaging and Communications in Medicine (DICOM) headers.

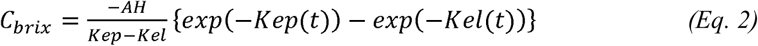

where C_brix_ is the concentration of CM in the tissue, K_el_ is the elimination constant of CM from the central compartment, K_ep_ is the exchange rate constant from the extracellular extravascular space (EES) to the plasma, and AH is the amplitude scaling constant [11].

### Proposed vascular-augmented composite model

In conventional PK modeling using the Brix model, myocardial tissue is represented as a two-compartment system comprising the plasma and EES. However, this approach assumes that the plasma compartment lies entirely outside the tissue voxel and does not explicitly account for intravascular signal contributions from the capillaries embedded within the myocardium.

This simplification may limit the accuracy of the model, particularly in organs such as the myocardium, where capillaries constitute a substantial portion of the tissue volume (approximately 20%) during mid-diastole□[12]. Therefore, a model that explicitly incorporates the vascular compartment within the myocardial voxel may accurately reflect the true distribution and kinetics of the contrast agents. To address this issue, we proposed a vascular-augmented composite model that represents the total myocardial contrast concentration as the sum of the intravascular and interstitial components, each weighted by its respective volume fractions within the voxel. A schematic representation of this model is shown in Figure 1.

**Figure 1.**
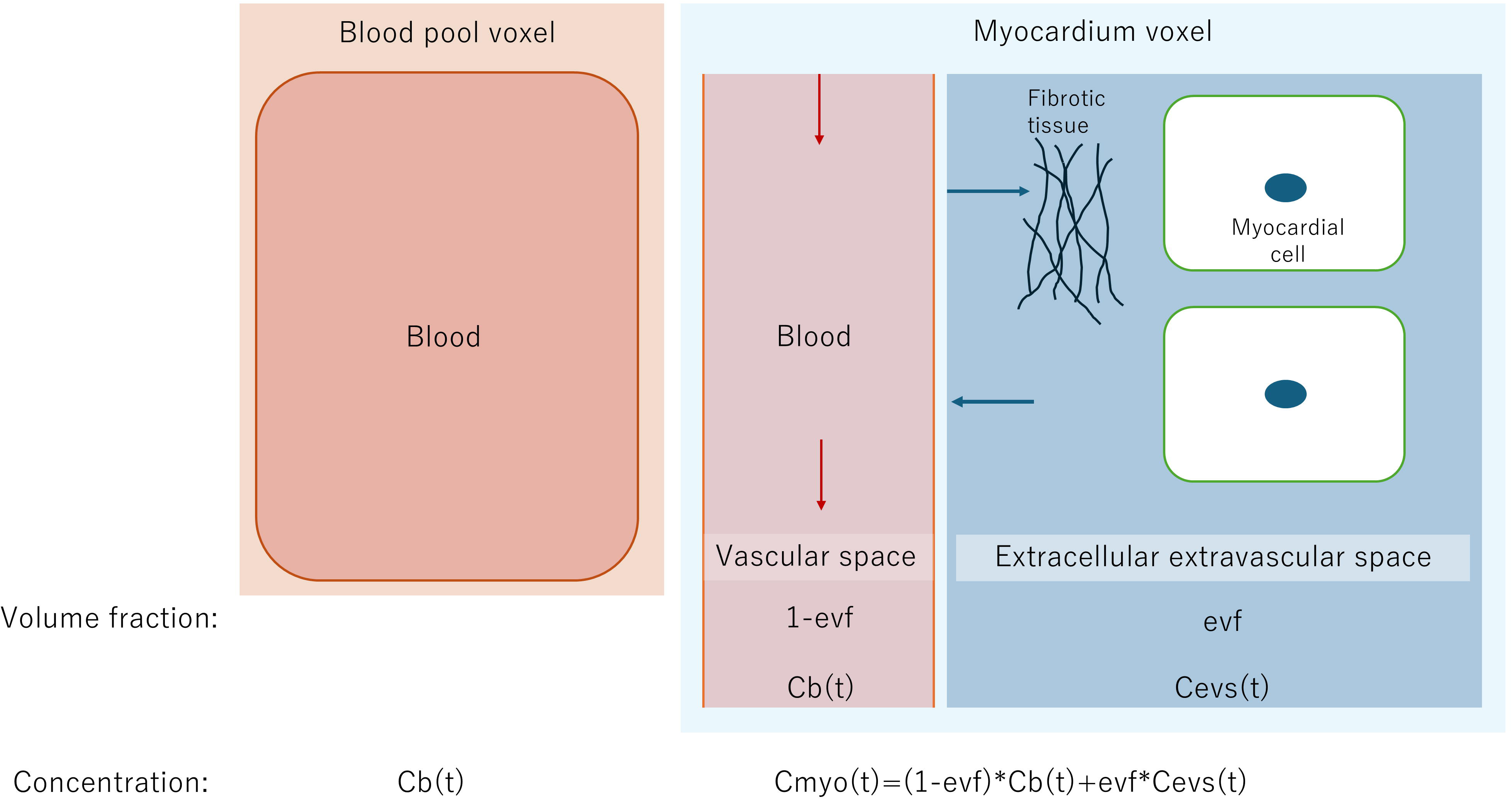
Schematic view of the composite model *C_b_(t)* contrast medium concentration in blood; *C_evs_(t)* contrast medium concentration in the extracellular extravascular space;, *evf,* extracellular extravascular space fraction; *C_myo_(t),* contrast medium concentration in the myocardium.

**Figure 2.**
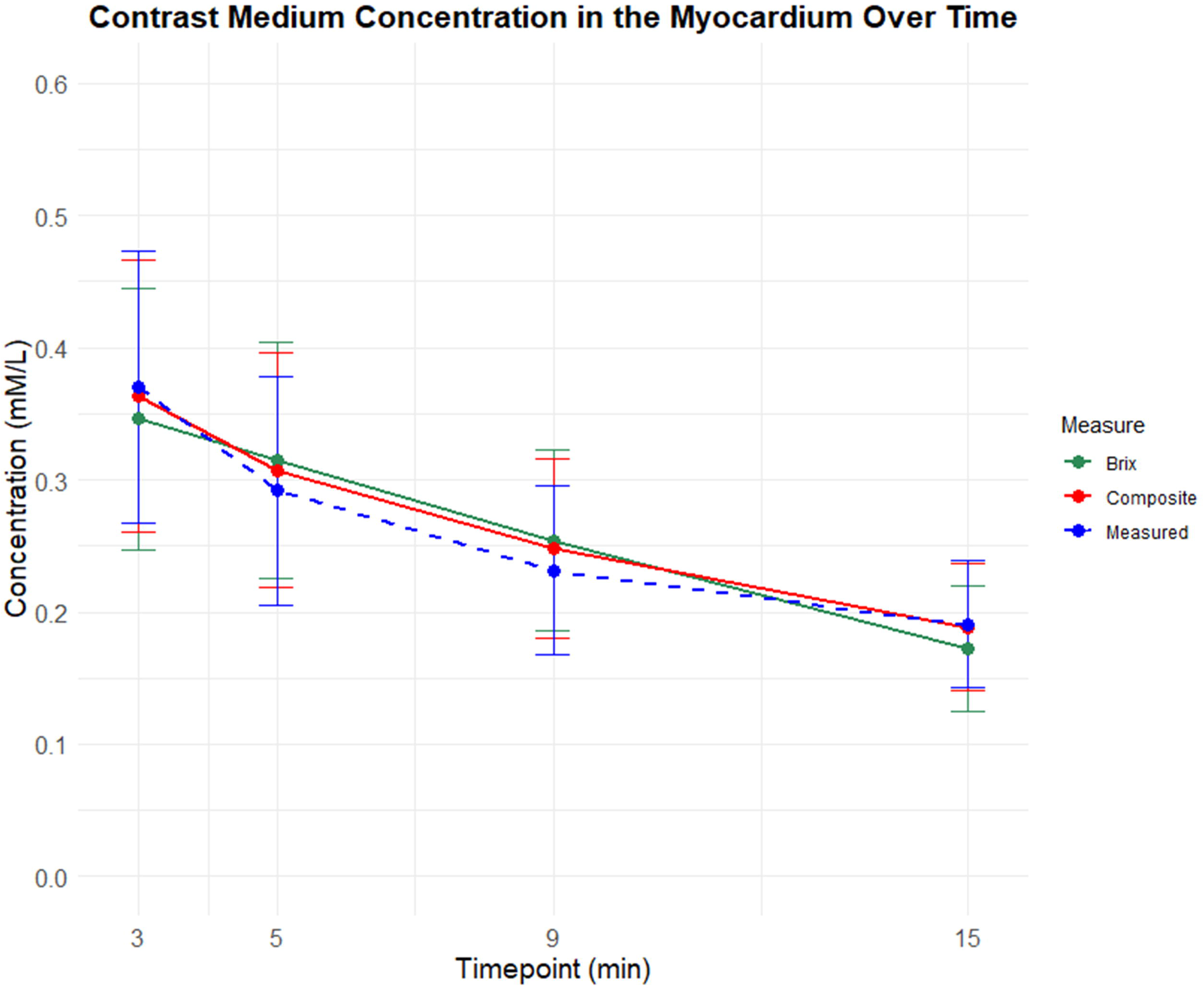
The line graph demonstrates the contrast medium concentration in the myocardium over time. The dotted lines indicate concentrations from the T1 maps. The myocardial contrast medium concentrations from the Brix model (green line) and composite model (red line) over time are also shown.

If evf represents the volume fraction of the EES within the myocardium and the remaining fraction (1 – evf) corresponds to the vascular compartment, the total myocardial concentration C(t) at time t is given by

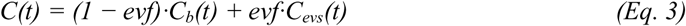

where C_b_(t) is the contrast concentration in the vascular space and C_evs_(t) is the contrast concentration in the EES.

The interstitial concentration, C_evs_(t), was modeled using the same formulation as the Brix model (Eq. 2). To model the vascular concentration C_b_(t), we used a bi-exponential function to approximate the wash-in and wash-out phases of contrast in the blood:

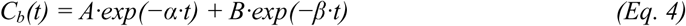

Where A and B are the scaling coefficients, and α and β are the distribution and elimination rate constants, respectively.

As CM was administered in two discrete boluses, the total concentration at time **t** must reflect the contributions of both doses [18]. Using the principle of superposition, the final myocardial concentration is

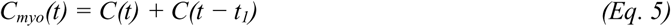

where *t* is the time interval between the first and second bolus injections.

The PK fitting of this model was performed using concentrations derived from four post-contrast T1 maps (2, 5, 9, and 15 min). All the models were implemented using Python 3.9 (http://www.python.org).

### Concentration analysis

After PK fitting, the myocardial CM concentrations at each time point were calculated from the measured and estimated concentrations from the PK models. Time points after contrast administration were collected from the DICOM tags of all post-contrast T1 maps, and concentrations at the same time were obtained from the PK model. The myocardial region was extracted by applying a threshold to the native T1map of each image [9]. For each participant, the myocardial pixel values from three slice images were averaged for comparison.

To examine the relationship between myocardial blood compartments and extracellular fluid fractions in myocardial tissue, we calculated the correlation between the mean myocardial blood compartment (1-evf) and the ECV value measured 15 min after CM administration using the PK model.

### Goodness of fit

The metrics used to compare the goodness of fit between the measured and estimated concentrations were the residual sum of squares (RSS), Akaike Information Criterion (AIC), and Bayesian Information Criterion (BIC).

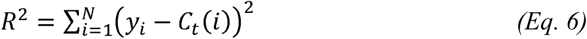

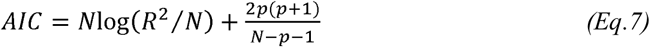

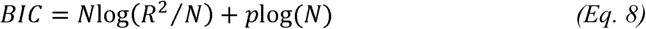

where, N is the number of observations, p is the number of model parameters, y_i_ is the observed value at time i, and C_t_(i) is the model estimate.

The RSS, AIC, and BIC of each pixel of the myocardium were calculated using both the Brix and composite methods.

### Statistical analysis

Statistical analyses were performed using the R statistical software (version 4.1.2; R Foundation for Statistical Computing). Normality was evaluated using the Shapiro–Wilk test. Continuous variables are expressed as mean□±standard deviation or as median (interquartile range), depending on the normality of distributions, and categorical variables as counts with percentages. Pearson’s correlation coefficient was used to evaluate the relationship between the myocardial blood fraction and ECV. Paired t-tests were used to compare the AIC, BIC, and the residual sum of squares between the methods. The prediction errors (| measured–estimated |) for each method were analyzed with a linear mixed□effects model (lme, REML package in R) including fixed effects for method, timepoint, and their interaction and a random intercept for subject; Type III analysis of variance tested the main and interaction effects, and post□hoc pairwise comparisons of marginal means were Bonferroni□adjusted. Model fit was compared across registration (no vs. co-registration) and time points by fitting simple linear regressions measured on estimated median concentrations and reporting the AIC and BIC. Statistical significance was set at P□<□0.05.

## Results

### Participants’ background

Participants’ background characteristics are shown in Table 1. Of the 107 participants (median age, 61 years), 71 (66%) were male. Median hematocrit and eGFR were 39% and 53 mL/min/1.73 m², respectively. Left ventricular functional parameters, including ejection fraction, volume, and mass, were representative of a clinically heterogeneous population.

**Table 1.**
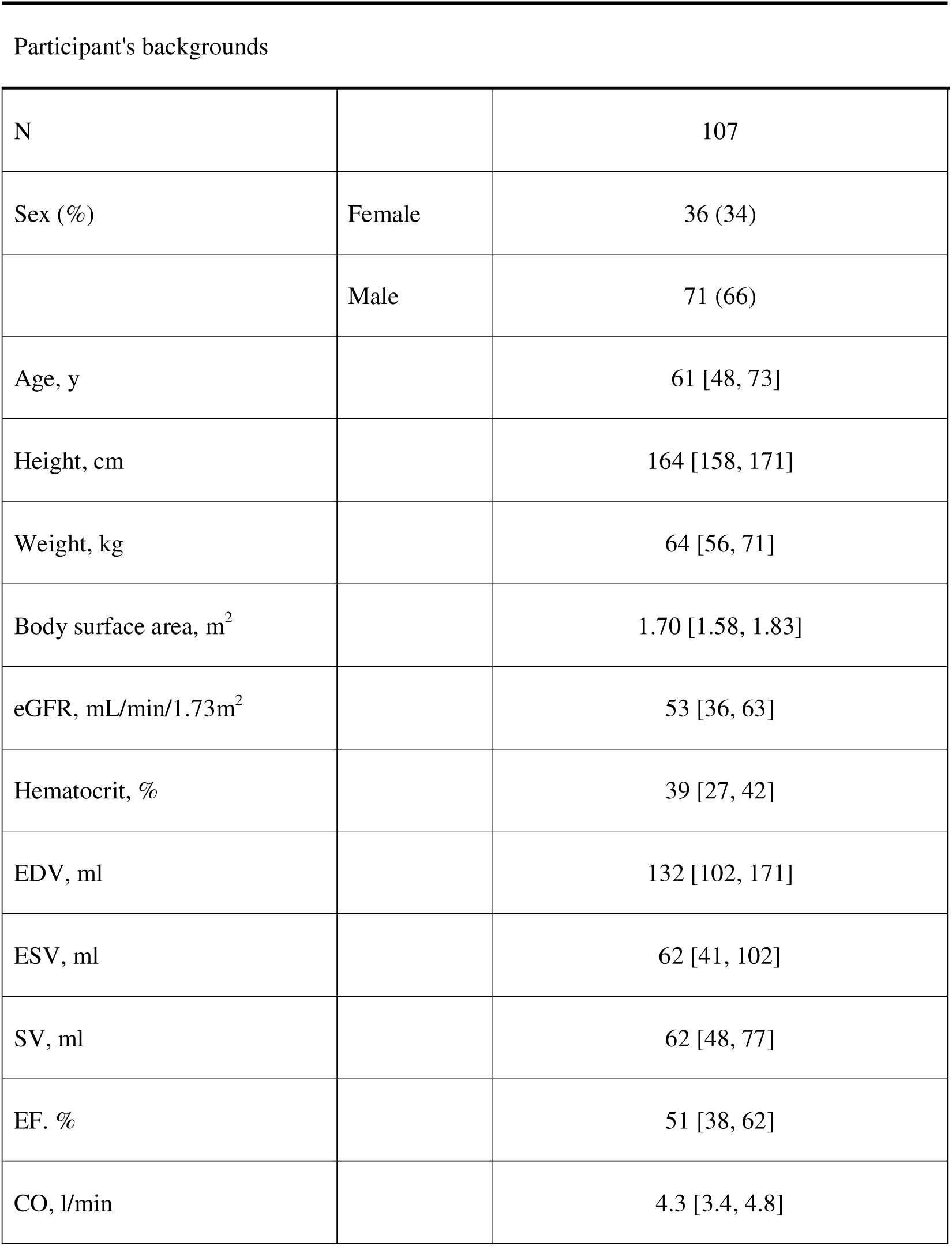
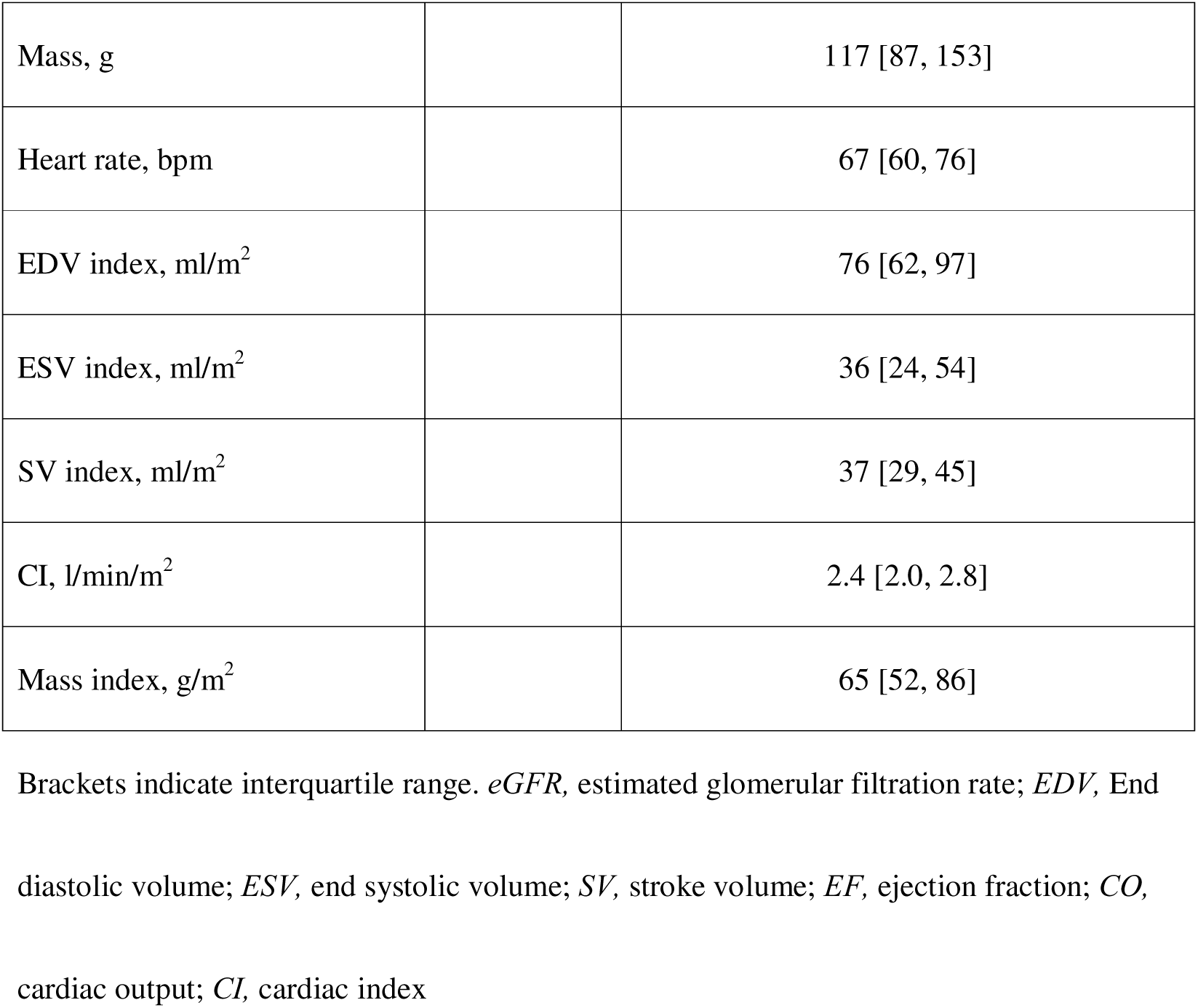

### Difference between measured and estimated concentrations

The estimated myocardial CM concentration calculated at each measurement time point demonstrated significant differences between the Brix and composite methods, with the composite model consistently yielding values closer to the measured concentrations (Table 2). Across all four time points, the mean absolute error of the composite model was consistently lower than that of the Brix model, indicating closer agreement with the reference (Table 2). Specifically, at 2□min, the difference between measured and estimated concentration was 0.007 for composite versus 0.024 for Brix; at 5□min, it was −0.016 vs. −0.023; at 9□min, −0.017 vs. −0.023; and at 15□min, 0.002 vs. 0.019. Paired t-tests confirmed that these differences were highly significant at all time point (Bonferroni-adjusted p□<□0.001). A linear mixed-effects model of absolute error (fixed effects: method, timepoint, and their interaction; random intercept per participant) revealed a significant main effect of method (F(1,440)=493.8, p□<□0.0001), a significant main effect of timepoint (F(3,440)=18.5, p□<□0.0001), and method×timepoint interaction (F(3,440)=25.0, p□<□0.0001). Post□hoc *emmeans* contrasts (Bonferroni-adjusted) demonstrated that at each time point, Brix errors exceeded composite errors by 0.0057–0.0146 (all p□<□0.0001), confirming the superior accuracy of the composite approach throughout the dynamic series. A line plot of the measured and estimated CM concentrations is shown in Figure 2.

**Table 2.**
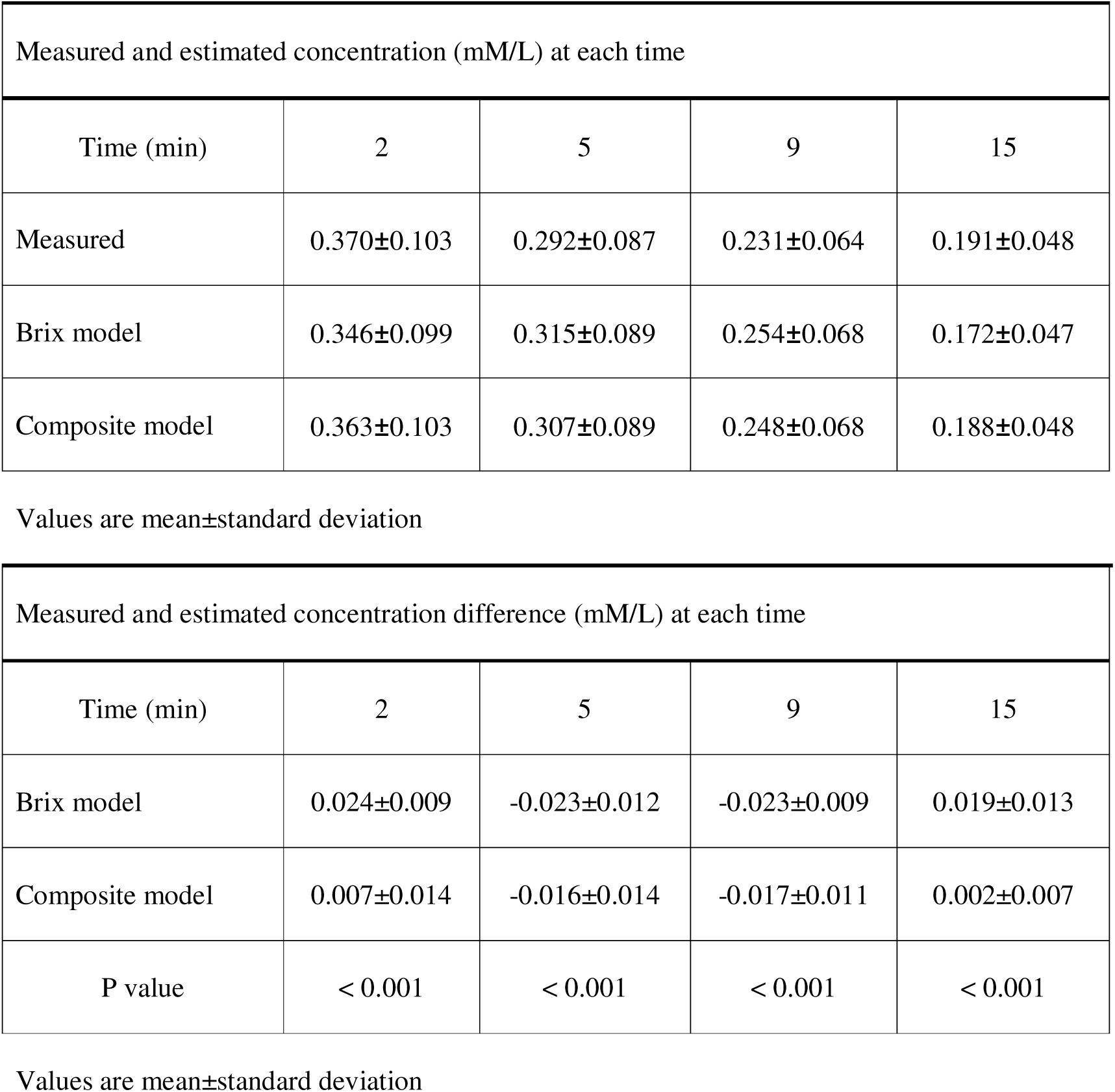

| Measured and estimated concentration (mM/L) at each time |  |  |  |  |
| --- | --- | --- | --- | --- |
| Time (min) | 2 | 5 | 9 | 15 |
| Measured | 0.370±0.103 | 0.292±0.087 | 0.231±0.064 | 0.191±0.048 |
| Brix model | 0.346±0.099 | 0.315±0.089 | 0.254±0.068 | 0.172±0.047 |
| Composite model | 0.363±0.103 | 0.307±0.089 | 0.248±0.068 | 0.188±0.048 |

Values are mean±standard deviation

| Measured and estimated concentration difference (mM/L) at each time |  |  |  |  |
| --- | --- | --- | --- | --- |
| Time (min) | 2 | 5 | 9 | 15 |
| Brix model | 0.024±0.009 | -0.023±0.012 | -0.023±0.009 | 0.019±0.013 |
| Composite model | 0.007±0.014 | -0.016±0.014 | -0.017±0.011 | 0.002±0.007 |
| P value | < 0.001 | < 0.001 | < 0.001 | < 0.001 |

Values are mean±standard deviation

The myocardial blood fraction in the analysis model and the ECV value from the T1 map were 35.0±12.9 and 34.1±8.68 (%), respectively. A positive correlation (r=0.61, p < 0.001) was found between the myocardial blood fraction (1-ecf) and ECV values measured from the T1 maps (Figure 3).

**Figure 3.**
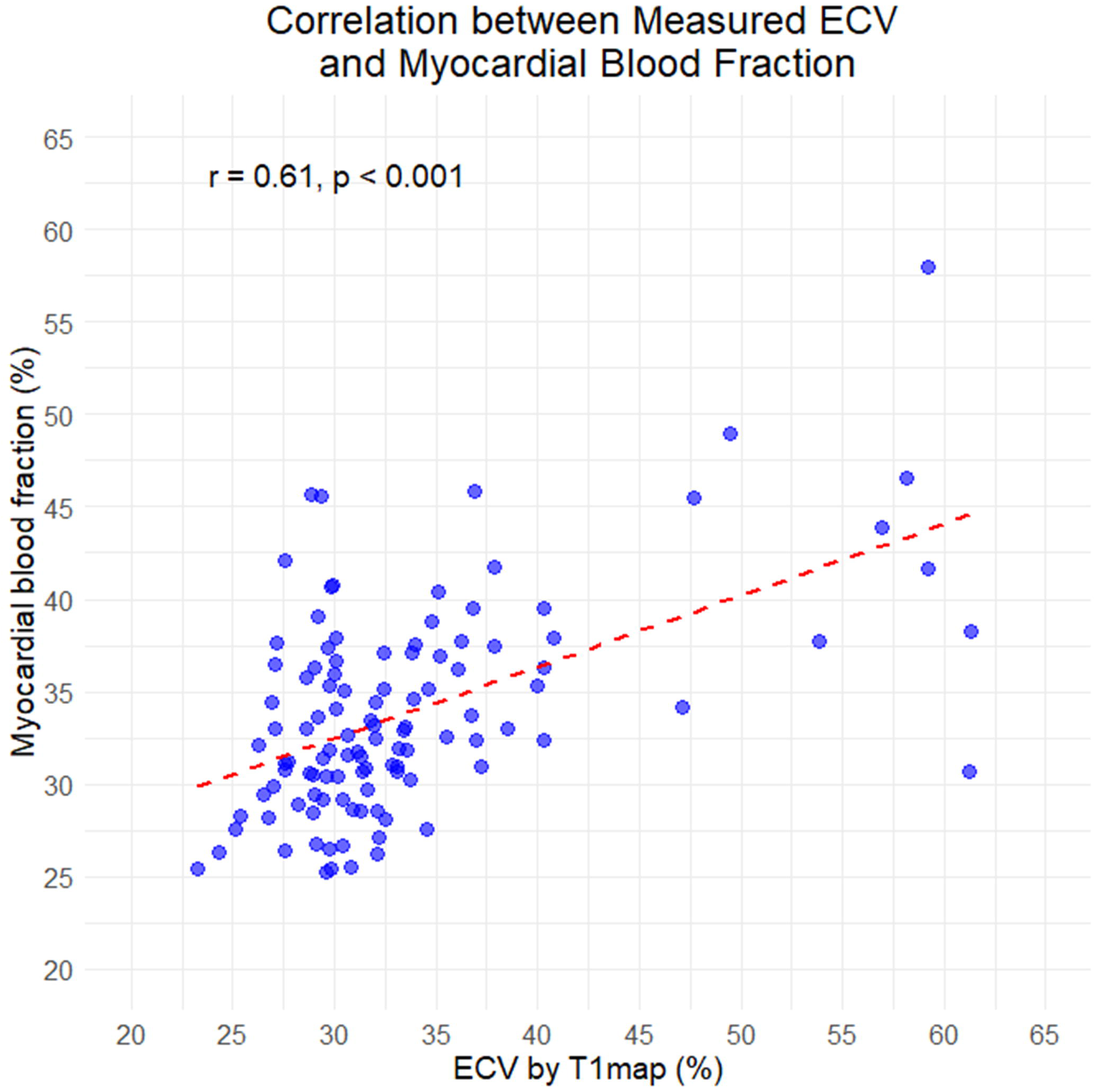
Correlation between the ECV from T1 map and myocardial blood fraction. A positive correlation (r=0.61, p < 0.001) was found between the two metrics. ECV, extracellular volume fraction

### Goodness of fit

The composite model demonstrated significantly lower AIC values compared to the Brix model (−71.13 ± 4.15 vs. −63.28 ± 3.43, p < 0.001) (Table 3). Similarly, the BIC values were significantly lower for the composite model than for the Brix model (−33.48 ± 4.01 vs. −25.62 ± 3.27, p < 0.001) (Table 3). The RSS was significantly lower in the composite model (0.00003 vs. 0.0007, p < 0.001) (Table 2). In 98.1% (105/107) of cases, both the AIC and BIC values were lower for the composite model than for the Brix model. The RSS was lower for the composite model in 83.2% (89/107) of cases. The estimated myocardial concentration and residual maps obtained from both the composite and Brix models are presented in Table 4. Representative concentration maps from the two models and their goodness of fit are shown in Figure 4.

**Figure 4.**
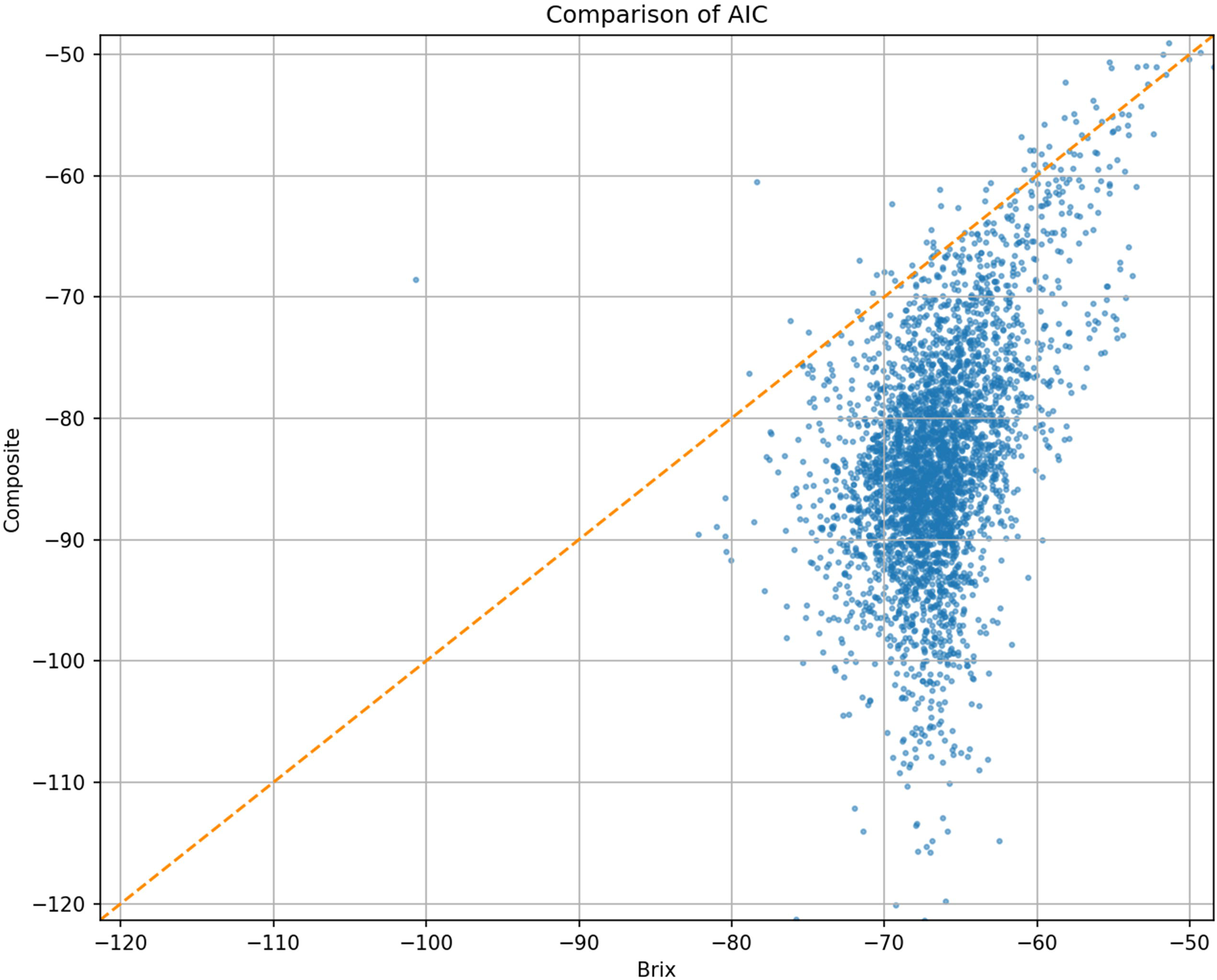
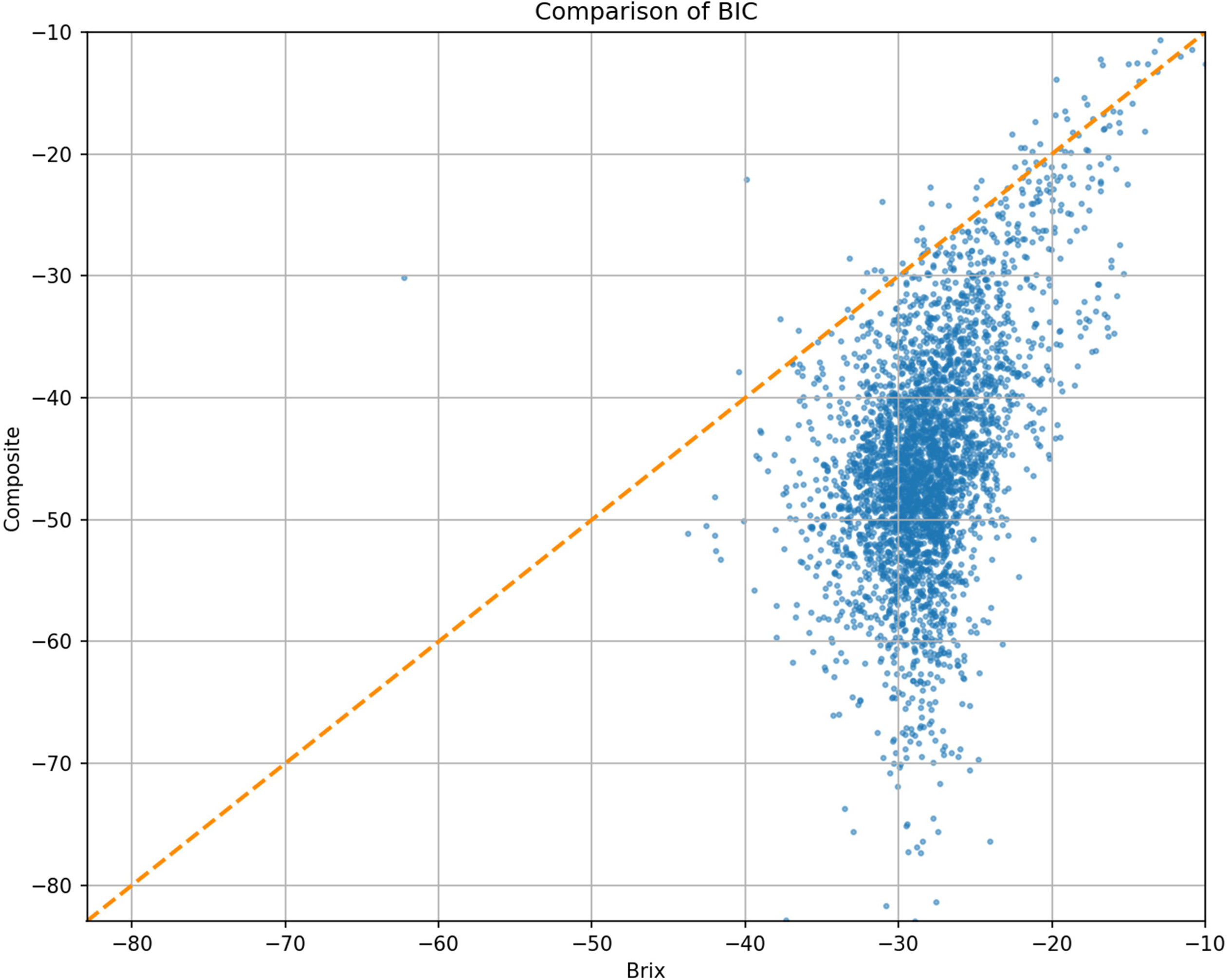
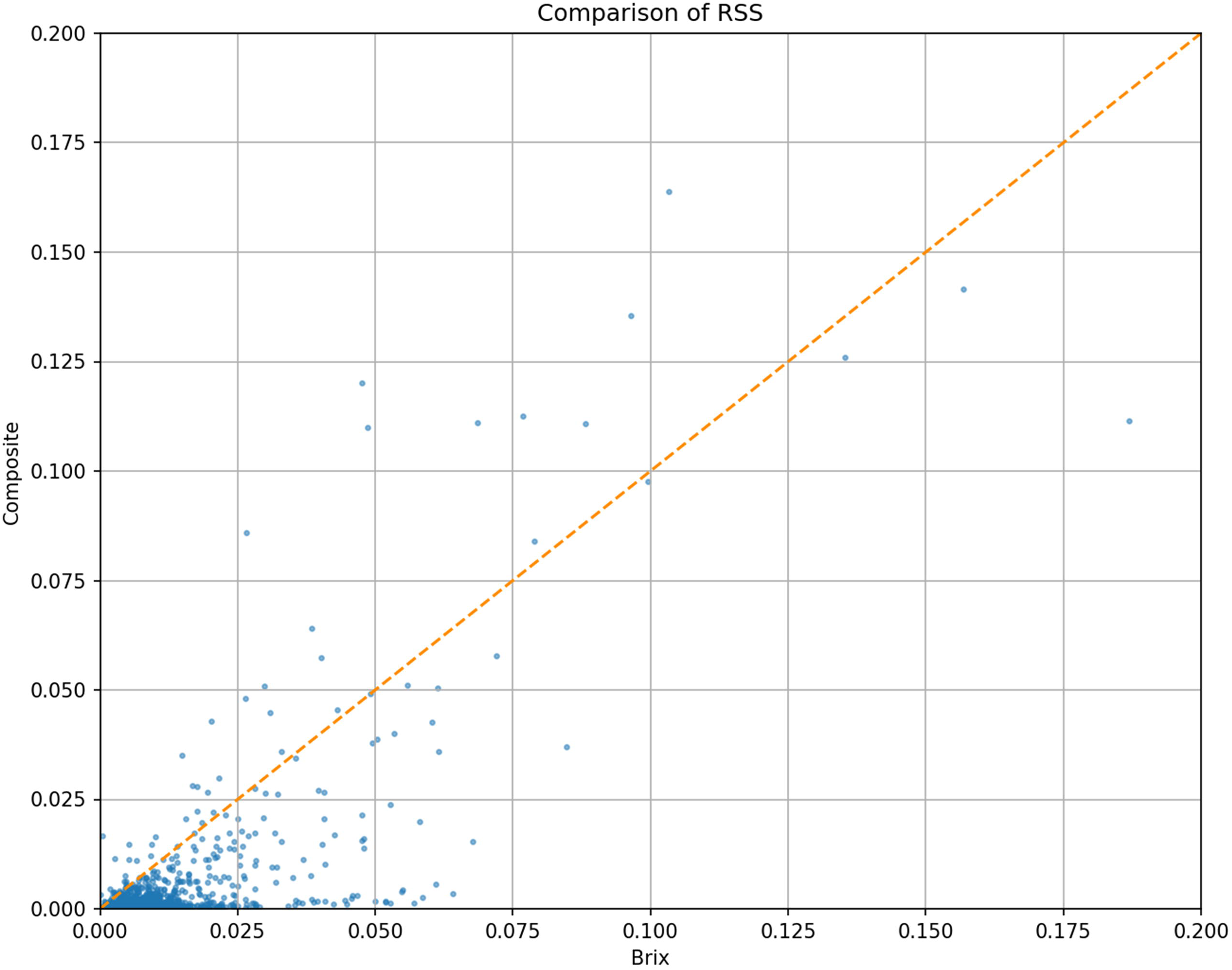
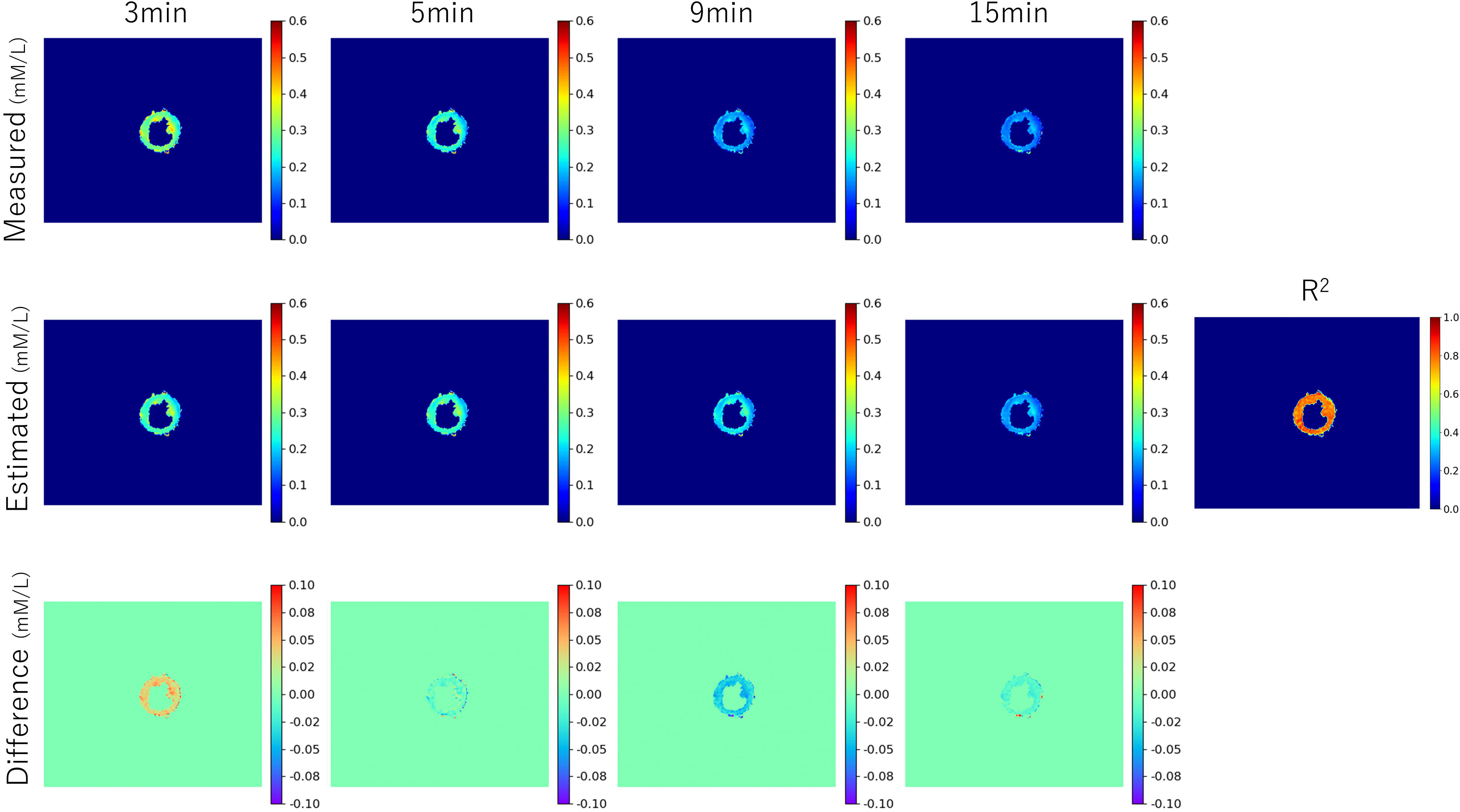
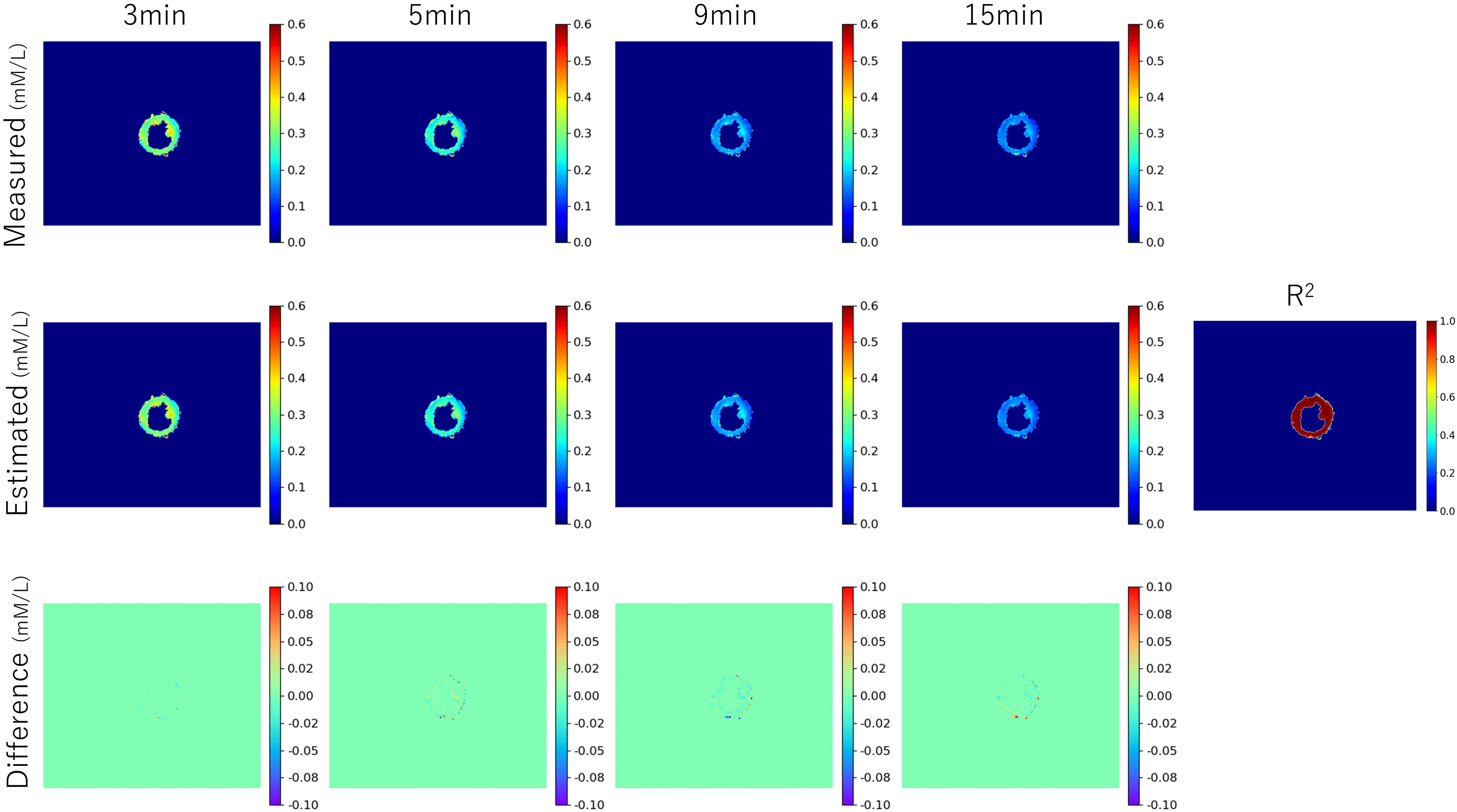
A case of a female with hypertrophic cardiomyopathy. The AICs of the Brix model and the composite model from each voxel are shown in (a). The average AICs for the Brix model and the composite model are −58.48 and −67.74, respectively. The BICs for the Brix and composite models from each voxel are displayed in (b). The mean BICs for the Brix and composite models are −20.93 and −30.19, respectively. The RSSs of the Brix and composite models from each voxel are presented in (c). The average RSSs for the Brix and composite models are 0.00023 and 0.000182, respectively. Measured and estimated myocardial contrast medium concentration maps, the difference between these maps, and the efficiency of determination (R^2^) map from the Brix model (d) and the composite model (e). The difference in myocardial contrast medium concentration between measured and estimated values is less (with less contrast to the background color) in the composite model. R^2^ shows a higher (redder color) value in the composite model. The R^2^ value is higher in the composite model. *AIC,* Akaike’s Information Criterion; *BIC,* Bayesian Information Criterion; *RSS,* residual sum of squares; *R^2^*, goodness of fit

**Table 3.**

| Goodness of fit for each model (n=107) |  | P value |
| --- | --- | --- |
| AIC by Brix | $-63.28 \pm 3.43$ | <0.001 |
| AIC by Composite | $-71.13 \pm 4.15$ | |
| BIC by Brix | $-25.62 \pm 3.27$ | <0.001 |
| BIC by Composite | $-33.48 \pm 4.01$ | |
| RSS by Brix | 0.00007 [0.00004, 0.00013] | <0.001 |
| RSS by Composite | 0.00003 [0.00002, 0.00009] |  |

Values are indicated in mean ± standard deviation or median with interquartile range. *AIC*, corrected Akaike information criterion; *BIC*, Bayesian information criterion; *RSS*, residual sum of squares

**Table 4.**

| The number of cases demonstrated a better fit of one model over another |  |  |
| --- | --- | --- |
| AIC: Composite < Brix | 105 | (98.1) |
| BIC: Composite < Brix | 105 | (98.1) |
| RSS: Composite < Brix | 89 | (83.2) |

Values are indicated in numbers (percentages). *AIC*, corrected Akaike information criterion; *BIC*, Bayesian information criterion; *RSS*, residual sum of squares.

## Discussion

The proposed vascular-augmented composite PK model, which incorporates myocardial vascular dynamics, demonstrated superior fitting accuracy to the measured T1-based contrast agent concentrations compared with the conventional Brix model at all time points.

Because myocardial tissue contains a substantial vascular component (approximately 22% in prior-micro-CT study [12]), we focused on the vascular regions within the extracellular vascular space of the tissue, which show similar dynamics to blood pools. Accordingly, a composite pharmacokinetic model was developed by incorporating an explicit vascular component [19] into the conventional Brix model [20], which describes contrast exchange between the blood pool and peripheral tissue compartments.

The myocardial blood space fraction averaged 0.34, indicating that a substantial proportion of myocardial tissue volume is attributable to the vascular compartment, demonstrating a positive correlation with the ECV. Although this value represents a model-based estimate rather than a direct measurement, it likely reflects the interstitial expansion within the myocardium. Because it correlates with the ECV, which encompasses the extracellular matrix and blood vessels, it is regarded as reflecting the interstitial space within the myocardium. The myocardial blood fraction is undercalculated in participants with high ECV; which predominantly corresponded to cases of cardiac amyloidosis. Pathological evaluation of the myocardium in amyloidosis demonstrated that amyloid deposits, coagulative necrosis, and fibrosis were observed in more than 61% of the extracellular space, and the number of small intramural coronary arteries did not increase [21], suggesting that the myocardial blood fraction is driven by pronounced interstitial expansion rather than increased vascular volume.

Regarding the significance of analyzing myocardial contrast washout, conventional myocardial tissue characterization using LGE imaging and ECV measurements is typically limited to a single post-contrast time point. ECV estimation assumes an equilibrium distribution of contrast between the blood pool and interstitium. While prior studies have reported no differences between the continuous infusion and single-dose contrast methods in normal myocardium, discrepancies have been observed in the pathological conditions associated with elevated ECV [22].

Although the variability in the migration speed of contrast agents across different tissues makes evaluation using conventional methods, such as LGE or T1 mapping-derived ECV, challenging within a single imaging session, dynamic assessment of myocardial contrast concentration enables longitudinal tissue analysis. This approach offers the potential for a more detailed *in vivo* tissue characterization. Several studies and simulations [23, 24] have reported findings that support this method.

A technical challenge in longitudinal evaluation is the need for acquiring images at the same time points across participants. In practice, precisely matching image acquisition times across participants can be challenging, and this issue has not been studied extensively. By applying a PK analysis model, evaluation times can be aligned across participants and an arbitrary evaluation time; however, this requires more precise model fitting.

When using the compartment model for tissue PK evaluation, as described in the methodology, a more accurate fit can be achieved by calculating the myocardial contrast agent concentration considering the difference in the dynamics between the tissue and blood regions in the ROI when the capillary volume in the myocardium is taken into account, as demonstrated in the present study. Data collection using dynamic T1 maps can be easily incorporated into existing protocols, including LGE imaging, because data are not collected continuously over a long period and images can be acquired with relatively high spatial resolution. The use of this approach enables more accurate modeling of tissue contrast agents.

This study had some limitations. First, a single MRI system was used in this single-center study. Second, these models are commonly used in DCE-MRI and, in theory, require at least 5–6 time points for time-series analysis. Reliable data measurements require sufficient time resolution (2–10 seconds) and measurements lasting at least 5 minutes [20]. In addition, these models require arterial input functions. The present method using T1 mapping cannot meet these requirements, and analysis using the Tofts model is challenging and cannot be performed. However, the strength of this method is that contrast agent concentrations can be quantified even in the absence of an AIF. Third, this study used T1 map-derived CM concentrations as the reference and could not compare them with the actual measured myocardial CM concentrations. Although this study is not directly comparable to other models, concepts from myocardial perfusion positron emission tomography were employed to improve model fitting.

## Conclusion

Upon comparison of kinetic models in PK modeling using myocardial dynamic T1 maps, the vascular-augmented composite model demonstrated improved goodness of fit compared with the Brix method, as it accounts for contrast agent dynamics within the myocardial vasculature.

## Ethics declarations Conflict of interest

The authors have no competing interests to declare that are relevant to the content of this article.

## Ethics approval

This study was performed in line with the principles of the Declaration of Helsinki. Approval was granted by the Ethics Committee of the National Cerebral and Cardiovascular Center (No. R20086). Written informed consent was obtained from all individual participants included in the study.

## Authors’ contributions

Yasutoshi Ohta: Acquisition of data, Study conception and design, Analysis and interpretation of data, Drafting of manuscript. Tomoro Morikawa: Acquisition of data, Formal analysis, Drafting of manuscript. Tatsuya Nishii: Acquisition of data, Drafting of manuscript. Yoshiaki Morita: Study conception and design, Drafting of manuscript. Tetsuya Fukuda: Study conception and design, Drafting of manuscript, Critical revision.

## Declaration of generative AI and AI-assisted technologies in the writing process

We declare that we have not used any generative AI or AI-assisted technologies in the writing process of this work.

## Data Availability

All data produced in the present study are available upon reasonable request to the authors.

## Acknowledgment

We would like to thank Editage (www.editage.jp) for English language editing.

## Data availability

The datasets for this study are not publicly available due to Japanese data protection laws.

## Finding

This work was supported by the Japan Society for the Promotion of Science (JSPS) KAKENHI Grant Number 22K15835.

## Acknowledgements

### Funding

This work was supported by JSPS KAKENHI grant number 22K15835.

### Data availability

The datasets and algorithms used and analyzed in the current study are available from the corresponding author upon reasonable request.

## References

1. Biglands JD, Ibraheem M, Magee DR, Radjenovic A, Plein S, Greenwood JP (2018) Quantitative myocardial perfusion imaging versus visual analysis in diagnosing myocardial ischemia: A CE-MARC substudy. JACC Cardiovasc Imaging 11:711–718.

2. Kim S, Loevner LA, Quon H, Kilger A, Sherman E, Weinstein G, Chalian A, Poptani H (2010) Prediction of response to chemoradiation therapy in squamous cell carcinomas of the head and neck using dynamic contrast-enhanced MR imaging. AJNR Am J Neuroradiol 31:262–268.

3. Taouli B, Johnson RS, Hajdu CH, Oei MTH, Merad M, Yee H, Rusinek H (2013) Hepatocellular carcinoma: perfusion quantification with dynamic contrast-enhanced MRI. AJR Am J Roentgenol 201:795–800.

4. Zheng X, Xiao L, Fan X, Huang N, Su Z, Xu X (2015) Free breathing DCE-MRI with motion correction and its values for benign and malignant liver tumor differentiation. Radiology of Infectious Diseases 2:65–71.

5. Maceira AM, Joshi J, Prasad SK, Moon JC, Perugini E, Harding I, Sheppard MN, Poole-Wilson PA, Hawkins PN, Pennell DJ (2005) Cardiovascular magnetic resonance in cardiac amyloidosis. Circulation 111:186–193.

6. Goldfarb JW, Zhao W (2014) Magnetic resonance imaging dynamic contrast enhancement (DCE) characteristics of healed myocardial infarction differ from viable myocardium. Magn Reson Imaging 32:1191–1197.

7. Chatzantonis G, Bietenbeck M, Florian A, Meier C, Stalling P, Korthals D, Reinecke H, Yilmaz A (2021) Diagnostic value of the novel CMR parameter “myocardial transit-time” (MyoTT) for the assessment of microvascular changes in cardiac amyloidosis and hypertrophic cardiomyopathy. Clin Res Cardiol 110:136–145.

8. Huang L-T, Zhang X, Li X, Malagi A, Huang Y, Guan X, Ho H, Kwan A, Wei J, Bi X, Christodoulou AG, Li D, Han H, Liu Y-W, Dharmakumar R, Yang H-J (2025) Improved myocardial tissue characterization using delayed-phase dynamic contrast-enhanced cardiac MRI. Radiol Cardiothorac Imaging 7:e240393.

9. Ohta Y, Shiotani M, Morita Y, Nishii T, Horinouchi H, Kotoku A, Fukuyama M, Tateishi E, Fukuda T (2025) Estimation of myocardial and blood gadolinium concentrations from T1 mapping via pharmacokinetic modeling: Influence of elastic deformation registration. medRxiv 2025.10.02.25337032.

10. Brix G, Semmler W, Port R, Schad LR, Layer G, Lorenz WJ (1991) Pharmacokinetic parameters in CNS Gd-DTPA enhanced MR imaging. J Comput Assist Tomogr 15:621–628.

11. Hoffmann U, Brix G, Knopp MV, Hess T, Lorenz WJ (1995) Pharmacokinetic mapping of the breast: a new method for dynamic MR mammography. Magn Reson Med 33:506–514.

12. Toyota E, Fujimoto K, Ogasawara Y, Kajita T, Shigeto F, Matsumoto T, Goto M, Kajiya F (2002) Dynamic changes in three-dimensional architecture and vascular volume of transmural coronary microvasculature between diastolic- and systolic-arrested rat hearts. Circulation 105:621–626.

13. Messroghli DR, Greiser A, Fröhlich M, Dietz R, Schulz-Menger J (2007) Optimization and validation of a fully-integrated pulse sequence for modified look-locker inversion-recovery (MOLLI) T1 mapping of the heart. J Magn Reson Imaging 26:1081–1086.

14. Kellman P, Wilson JR, Xue H, Ugander M, Arai AE (2012) Extracellular volume fraction mapping in the myocardium, part 1: evaluation of an automated method. J Cardiovasc Magn Reson 14:63.

15. Xue H, Shah S, Greiser A, Guetter C, Littmann A, Jolly MP, Arai AE, Zuehlsdorff S, Guehring J, Kellman P (2012) Motion correction for myocardial T1 mapping using image registration with synthetic image estimation. Magn Reson Med 67:1644–1655.

16. (2024) ACR Manual on Contrast Media. https://www.acr.org/-/media/acr/files/clinical-resources/contrast_media.pdf. Accessed 8 Sept 2024

17. Sanvito F, Raymond C, Cho NS, Yao J, Hagiwara A, Orpilla J, Liau LM, Everson RG, Nghiemphu PL, Lai A, Prins R, Salamon N, Cloughesy TF, Ellingson BM (2023) Simultaneous quantification of perfusion, permeability, and leakage effects in brain gliomas using dynamic spin-and-gradient-echo echoplanar imaging MRI. Eur Radiol. doi: 10.1007/s00330-023-10215-z

18. Wang W, Ouyang SP (1998) The formulation of the principle of superposition in the presence of non-compliance and its applications in multiple dose pharmacokinetics. J Pharmacokinet Biopharm 26:457–469.

19. Yoshida K, Mullani N, Gould KL (1996) Coronary flow and flow reserve by PET simplified for clinical applications using rubidium-82 or nitrogen-13-ammonia. J Nucl Med 37:1701–1712.

20. Tofts PS, Brix G, Buckley DL, Evelhoch JL, Henderson E, Knopp MV, Larsson HB, Lee TY, Mayr NA, Parker GJ, Port RE, Taylor J, Weisskoff RM (1999) Estimating kinetic parameters from dynamic contrast-enhanced T(1)-weighted MRI of a diffusable tracer: standardized quantities and symbols. J Magn Reson Imaging 10:223–232.

21. Hashimura H, Ishibashi-Ueda H, Yonemoto Y, Ohta-Ogo K, Matsuyama T-A, Ikeda Y, Morita Y, Yamada N, Yasui H, Naito H (2016) Late gadolinium enhancement in cardiac amyloidosis: attributable both to interstitial amyloid deposition and subendocardial fibrosis caused by ischemia. Heart Vessels 31:990–995.

22. White SK, Sado DM, Fontana M, Banypersad SM, Maestrini V, Flett AS, Piechnik SK, Robson MD, Hausenloy DJ, Sheikh AM, Hawkins PN, Moon JC (2013) T1 mapping for myocardial extracellular volume measurement by CMR: bolus only versus primed infusion technique. JACC Cardiovasc Imaging 6:955–962.

23. Axel L (2023) Modeling of factors affecting late gadolinium enhancement kinetics in MRI of cardiac amyloid. J Cardiovasc Magn Reson 25:46.

24. Moran GR, Thornhill RE, Sykes J, Prato FS (2002) Myocardial viability imaging using Gd-DTPA: physiological modeling of infarcted myocardium, and impact on injection strategy and imaging time. Magn Reson Med 48:791–800.

