## Supplemental material for "Vascular-Augmented Two-Compartment Fitting Improves Model Performance for Intermittent Myocardial T1 Mapping"

**Supplemental Table 1**

| Participant's backgrounds |  |  |
| --- | --- | --- |
| n |  | 107 |
| Sex (%) | Female | 36 (34) |
|  | Male | 71 (66) |
| Age, yrs |  | 61 [48, 73] |
| Height, cm |  | 164 [158, 171] |
| Weight, kg |  | 64 [56, 71] |
| Body surface area, m^2^ |  | 1.70 [1.58, 1.83] |
| eGFR, mL/min/1.73m^2^ |  | 53 [36, 63] |
| Hematocrit, % |  | 39 [27, 42] |
| EDV, ml |  | 132 [102, 171] |
| ESV, ml |  | 62 [41, 102] |
| SV, ml |  | 62 [48, 77] |
| EF. % |  | 51 [38, 62] |
| CO, l/min |  | 4.3 [3.4, 4.8] |
| Mass, g |  | 117 [87, 153] |
| Heart rate, bpm |  | 67 [60, 76] |
| EDV index, ml/m^2^ |  | 76 [62, 97] |
| ESV index, ml/m^2^ |  | 36 [24, 54] |
| SV index, ml/m^2^ |  | 37 [29, 45] |
| CI, l/min/m^2^ |  | 2.4 [2.0, 2.8] |
| Mass index, g/m^2^ |  | 65 [52, 86] |
| Brackets indicate interquartile range. *eGFR* estimated glomerular filtration rate, *EDV* End diastolic volume, *ESV* end systolic volume, *SV* stroke volume, *EF* ejection fraction, *CO* cardiac output, *CI* cardiac index | | |
